# Transcriptomic-based clustering of advanced atherosclerotic plaques identifies subgroups of plaques with differential underlying biology that associate with clinical presentation

**DOI:** 10.1101/2021.11.25.21266855

**Authors:** Michal Mokry, Arjan Boltjes, Kai Cui, Lotte Slenders, Joost M. Mekke, Marie A.C. Depuydt, Nathalie Timmerman, Farahnaz Waissi, Maarten C Verwer, Adam W. Turner, Mohammad Daud Khan, Chani J. Hodonsky, Ernest Diez Benavente, Robin J.G. Hartman, Noortje A M van den Dungen, Nico Lansu, Emilia Nagyova, Koen H.M. Prange, Eleftherios Pavlos, Evangelos Andreakos, Heribert Schunkert, Gary K. Owens, Claudia Monaco, Aloke V Finn, Renu Virmani, Nicholas J. Leeper, Menno P.J. de Winther, Johan Kuiper, Gert J. de Borst, Erik S.G. Stroes, Mete Civelek, Dominique P.V. de Kleijn, Hester M. den Ruijter, Folkert W. Asselbergs, Sander W. van der Laan, Clint L. Miller, Gerard Pasterkamp

## Abstract

Histopathological studies have revealed key processes of atherosclerotic plaque thrombosis. However, the diversity and complexity of lesion types highlight the need for improved sub- phenotyping. We hypothesized that unbiased clustering of plaques based on gene expression results in an alternative categorization of late-stage atherosclerotic lesions.

We analyzed the gene expression profiles of 654 advanced human carotid plaques. The unsupervised, transcriptome-driven clustering revealed five dominant plaque types. These novel plaque phenotypes associated with clinical presentation (p<0.001) and showed differences in cellular compositions. Validation in coronary segments showed that the molecular signature of these plaques was linked to coronary ischemia. One of the plaque types with most severe clinical symptoms pointed to both inflammatory and fibrotic cell lineages. This highlighted plaque phenotype showed high expression of genes involved in active inflammatory processes, neutrophil degranulation, matrix turnover, and metabolism. For clinical translation, we did a first promising attempt to identify circulating biomarkers that mark these newly identified plaque phenotypes.

In conclusion, the definition of the plaque at risk for a thrombotic event can be fine-tuned by in-depth transcriptomic based phenotyping. These differential plaque phenotypes prove clinically relevant for both carotid and coronary artery plaques and point to differential underlying biology of symptomatic lesions.

## Introduction

The classical concept of the ‘vulnerable plaque’ that depicts the plaque rupture as the major pathological substrate for acute cardiovascular events originated in the 1980s from observations in patients who died of coronary syndromes^1, 2^. This recognition spawned a generation of research that led to a greater understanding of how complicated atherosclerotic plaques form and precipitate into thromboembolic events secondary to plaque rupture. Current evidence suggests that a sole focus on plaque rupture of atheromatous lesions in clinical and basic research may have oversimplified the complex collection of atherosclerotic diseases and obscured other mechanisms that may mandate different management strategies.

In addition to histology, molecular phenotyping using the whole transcriptome provides more resolution and allows an in-depth understanding and discovery of processes active in the diseased vascular tissue^3–6^. These studies successfully utilized the gene co-expression networks or compared cases and controls. However, they did not attempt to redefine the plaque type definitions based on gene expression signatures. Unsupervised cluster analysis, based on transcriptomic datasets, can be used to group patients with similar molecular characteristics of the diseased tissue and have the potential to unravel disease phenotypes that fine-tunes the patho-histological evaluation^7–10^. This approach is often used in cancer research and led to the identification of novel tumor subtypes. We, therefore hypothesized that unbiased clustering based on gene expression of human advanced atherosclerotic plaques unveils novel phenotypes of late-stage human atherosclerotic plaques.

Using a multi-layered approach, we created gene expression maps within a large biobank of advanced carotid lesions (n=654) and studied histological characteristics and the clinical presentation. We report that transcriptome-based analysis of human atherosclerotic lesions identified five plaque clusters linked to the occurrence of clinical events and biological processes. We highlight a plaque type that is enriched with *ACTA2* as well as *CD14* expressing cells and with the highest expression of genes involved in neutrophil degranulation, mTOR, iron uptake, and other pathways linked to active inflammatory response and increased expression of genes involved in glycolysis. We verified these findings in coronary artery segments where this plaque type predicted coronary ischemic events. Finally, we did a first pilot study in search for circulating biomarkers that reflect these transcriptomic clusters. Our data demonstrate that a transcriptome based plaque characterization may have significant added value in phenotyping advanced atherosclerotic lesions that lead to clinical symptoms.

## Results

### Athero-Express study cohort of carotid segments

After quality control, based on measures of RNA library complexity, we have used 654 patients for clustering analysis. From those patients, the accessibility of clinical data resulted in the inclusion of 632 plaques for the prediction models. The baseline characteristics of patients from Athero-Express selected for this plaque study are provided in Supplementary Table 1. In total, 75.3% of included patients were males, 24.7% were females with a mean age of 68.4 years and a mean BMI of 26.6. 43.2% of included patients had a stroke, 24.2% transient ischemic attack, 17.3% of patients had ocular symptoms, and the remaining 15.3% were asymptomatic. In 3-year follow up 13.1% of patients suffered major adverse cardiovascular events (MACE). Histologically, 30.0% of included plaques were classified as atheromatous, 32.3% as fibrous, and 37.7% as fibro-atheromatous.

### Transcriptome-defined molecular plaque types

To identify groups of patients with similar molecular signatures of advanced atherosclerotic lesion characteristics, we have utilized 654 individual transcriptomes from plaques (Fig. 1a) that passed the QC filters and identified five major molecular plaque types - referred to as #0, #1, #2, #3 and #4 (Fig. 1b, Extended Data figure 1ab). All five clusters contain samples with similar numbers of detected protein-coding genes (Extended Data figure 1c). Based on the principal component analysis (PCA) projection and correlation to the most similar sample (Extended Data figure 1c), type #3 contains more heterogeneous plaques while the other clusters demonstrate higher correlations between the samples. Overall, we did not observe clear boundaries between five plaque types, suggesting the presence of intermediate types (Fig. 1b, Extended Data figure 1ab).

**Figure 1.**
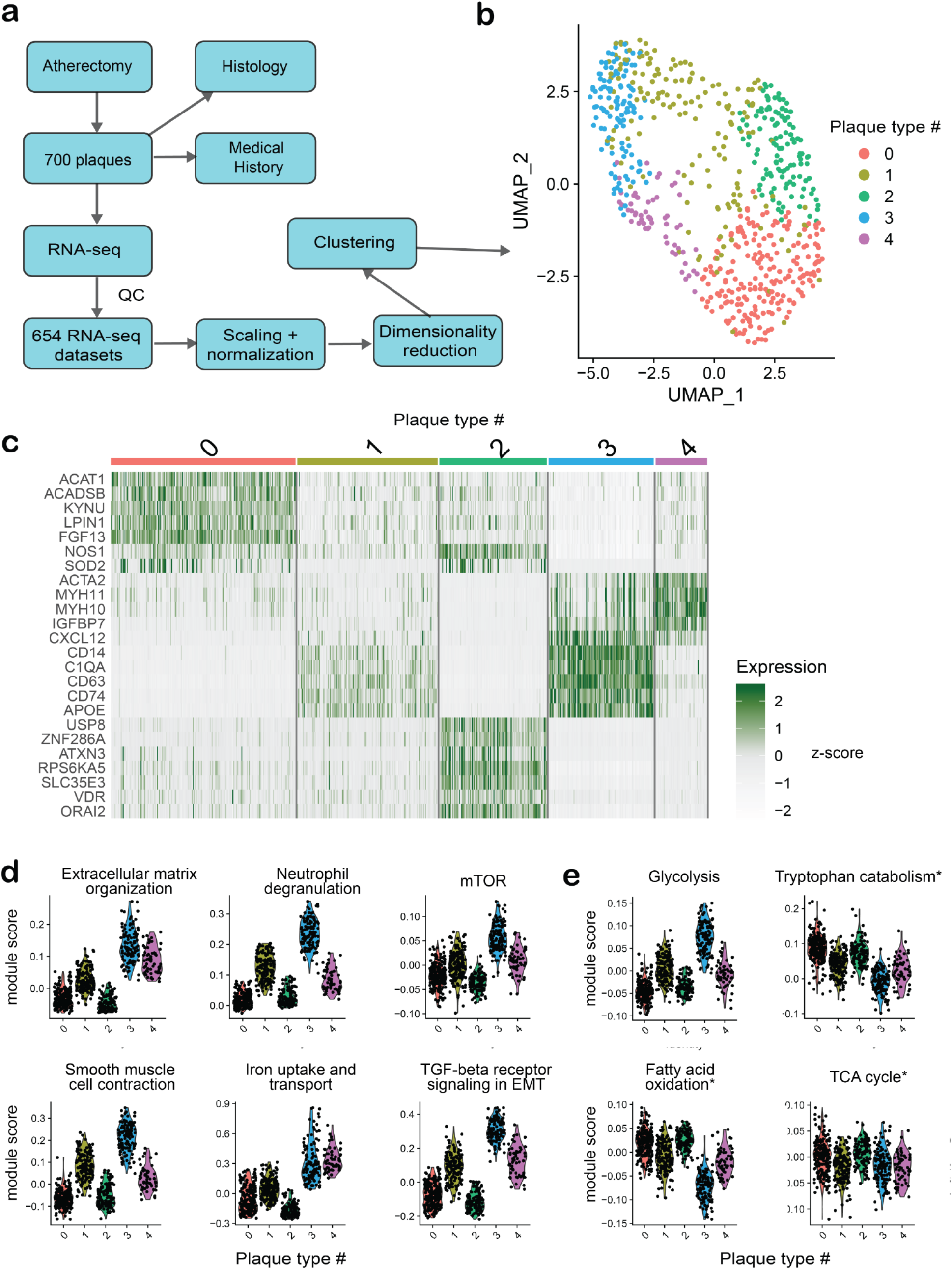
Unsupervised clustering of plaques based on transcriptomics data. a) A schematic workflow of the clustering analysis. b) UMAP projection of the 654 plaque samples based on RNA-seq dataset. The color indicates the cluster corresponding to the plaque type from the SNN modularity optimization-based clustering algorithm. c) Heatmap depicts relative gene expression levels of selected plaque type enriched genes in individual samples and plaque clusters. d) Module scores of genes annotated to selected molecular pathways in different plaque clusters. e) Module scores of genes annotated to selected metabolic pathways in different plaque clusters. * depicts empirically selected pathways that were not significantly enriched in the pathway analysis.

The five transcriptomics-based plaque types vastly differ in the expression of numerous candidate marker genes (Fig. 1c, Supplementary Table 2) what supports that the clustering is robust and reflects different underlying biology. For instance, type #0 plaques show increased expression of *FGF13*, *LPIN1*, and *KYNU*. Type #1 and #3 have increased expression of inflammatory molecules and leukocyte markers (*CXCL12*, *C1QA*, *CD14*, *CD73*, or *APOE*), while #3 also shows a modestly increased expression of classical smooth muscle cell markers (*MYH11*, *MYH10*, and *ACTA2*). Type #2 plaques show increased expression of *NOS1*, *SOD2*, *VDR*, *SLC35E3*, and *ATXN3*, while they mostly lack expression of immune cells and SMC markers. Finally, type #4 has the highest expression of classical smooth muscle cell markers (MYH11, MYH10, and ACTA2) and reduced inflammatory and leukocyte marker genes expression.

Next, to understand which molecular processes and pathways underlie the five molecular plaque types, we performed pathway analysis on genes upregulated (based on the differential gene expression analysis) in the individual clusters. Clusters # 1, #3, and #4 showed significant enrichment in numerous pathways (Supplementary Table 3). Increased expression of genes involved in neutrophil degranulation, mTOR, iron uptake, and other pathways linked to active inflammatory-response-related processes overlapped with increased expression of glycolysis genes, specifically in type #3 (Fig. 1de). The same plaque type showed decreased activity of genes involved in fatty acids oxidation, while the activity of the citric acid cycle (TCA cycle) seems to be comparable in all clusters. Similarly, the processes involved in extracellular matrix homeostasis (synthesis, organization, degradation) and cell plasticity (endothelial-mesenchymal transition - EMT) showed specific enrichment in individual plaque types.

The genes upregulated in plaque types #0 and #2 did not show significant enrichment in the pathway analysis. However, the projection of pathways that involve some of the cluster-specific genes (for example, the tryptophan catabolism, which involves L-kynureninase - *KYNU*) showed increased expression in these specific clusters.

Altogether, the transcriptome-defined plaque types differ in molecular signatures involving metabolism, the inflammatory response, and the process involved in extracellular matrix (ECM) homeostasis. This suggest that the clustering is robust and driven by different underlying biology

### Transcriptome-defined molecular plaque types differ in histological and cellular composition

Of interest, one specific plaque type (#3) was characterized by gene enrichment pointing to cell types and processes that are commonly used to differentiate between stable and unstable plaques. This type #3 showed high expression of genes specific for inflammatory cells (like C1QA and CD14) and cells responsible for a fibrotic phenotype (*MYH11* and *ACTA2*). Since the variation in the cellular composition of complex tissues is one of the main drivers of transcriptomic differences, we have analyzed the histological features of the five molecular plaque clusters (Fig 2a). Clusters #0 and #4 were significantly enriched in fibrous plaques with lower fat content and increased expression for *ACTA2*. On the contrary, type #2 was more enriched in atheromatous plaques, high-fat content, and increased presence of *CD68* positive cells. For clusters #0 to #4 there was no distinct pattern for the presence of calcification, media remnants, collagen content, and intraplaque hemorrhage (IPH).

**Figure 2.**
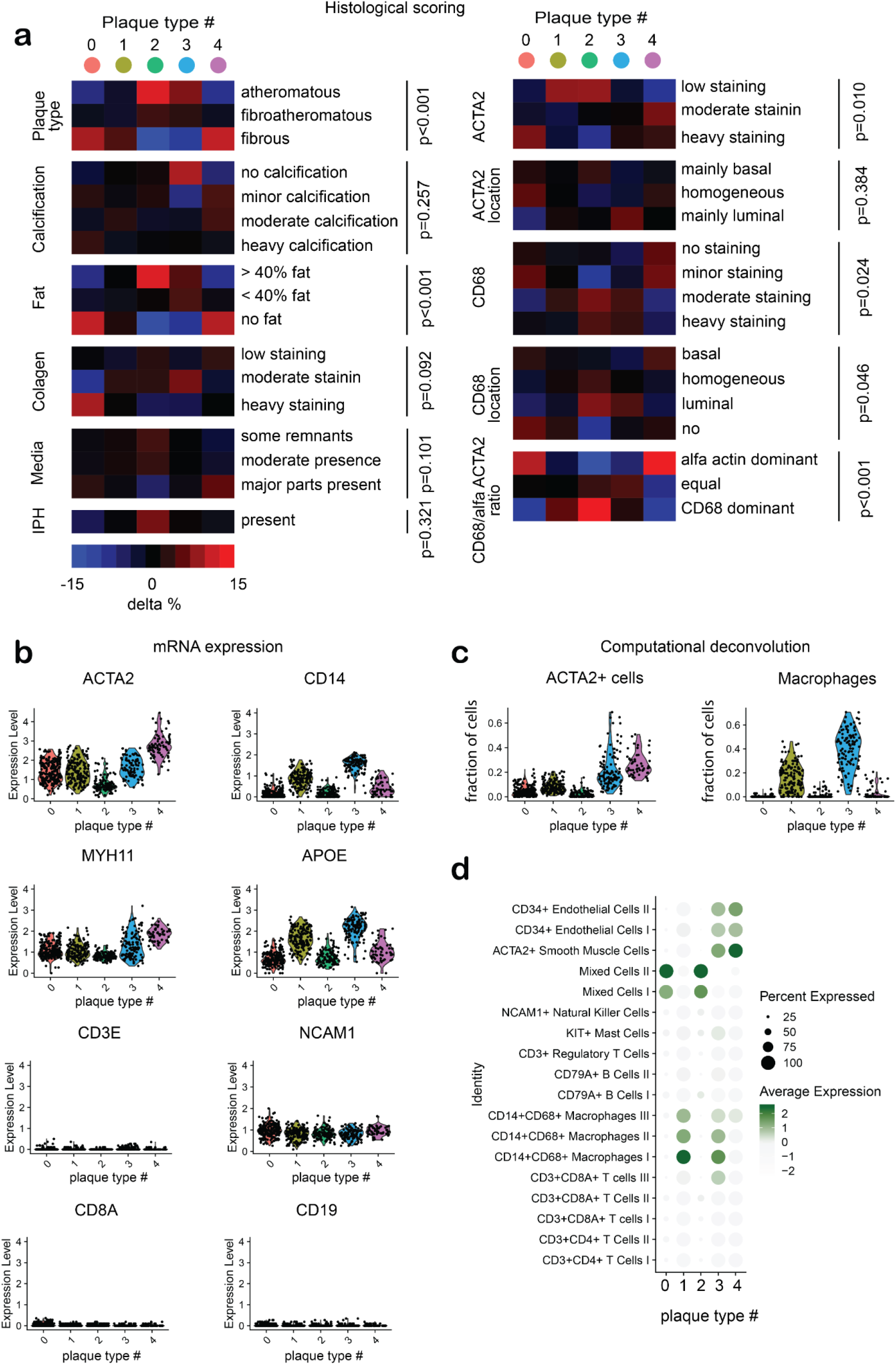
Unsupervised clustering of plaques based and cell composition. a) Distribution of histological features in five transcriptome-based clusters. The percentage represents the difference over equal distribution. (IPH - intraplaque hemorrhage). P-values were calculated using the chi-square test for categorical variables and one-way analysis of variance (ANOVA) for continuous variables. b) Expression of selected cell marker genes in transcriptome-based clusters. c) Fraction of specific cell types contributing to the bulk RNA-seq signal deconvoluted using the reference-based on single-cell transcriptomics dataset. d) Expression of cluster-specific gene sets in single-cell transcriptomics datasets from atherosclerotic plaques.

Even though the plaque clusters #0 and # 4 showed both absolute and relative increase in *ACTA2* positive cells in the histological evaluation, only in plaque-type #4 was this followed by upregulation of classical SMC markers like *ACTA2* or *MYH11* in the transcriptomic data (Fig 2b). Similarly, genes involved in ECM organization were downregulated in type #0 compared to #4 and #3.

Since the presence of a specific cell type does not necessarily correlate with individual marker genes, we utilized carotid lesion single-cell RNA-seq based deconvolution (MuSiC algorithm^11^), to estimate the relative cell composition of each plaque sample (Fig. 2c). In line with the marker gene expression, plaque clusters #3 and #4 showed a higher percentage of *ACTA2*+ cells relative to type #0. In line with cell deconvolution, genes overexpressed in type #0 did not show specific expression in *ACTA2*+ cells in single-cell transcriptomics data (Fig. 2d); instead, they seemed to be expressed in populations identified as “mixed cells” - a population without a clear cell type–defining expression profile. This mixed plaque type seemed to contain apoptotic myeloid and T cells^12^ and express some of the foam cell drivers (*LRP1B*^13^). Altogether, the histologically scored numbers of *ACTA2*+ cells in plaque type #0, which resides in a highly fibrotic environment, seems to be overestimated or represents a transcriptionally less active population.

Similarly, the histological evaluation of plaque type #2 showed the association with increased *CD68*+ cells. However, the expression of macrophage markers like *CD14* and genes involved in inflammatory pathways was predominantly downregulated at the RNA level. Plaques from cluster #2 also show a lower percentage of macrophages in computationally deconvoluted data. Besides, these cluster-specific genes project to a likely apoptotic “mixed cell” population. It suggests that the residing macrophages in type #2 plaques have decreased transcriptional activity, or their numbers in the histological evaluation are overestimated. Notably, plaque types do not seem to be driven by the variation in NK-, B- or T-cells content (Fig 2bd).

Together, by combining transcriptomics and histological data, we categorized the five plaque clusters as follows: #0 – fibro-collagenous, #1 - intermediate, #2 - lipomatous, #3 - fibro-inflammatory, and #4 fibro- cellular.

### Transcriptome-defined molecular plaque types reflect disease severity

Next, to gain clinical insights into the distinct transcriptome-based plaque clusters, we explored the severity of clinical symptoms before the surgery. The clusters #2, #3 revealed higher percentages of more severe symptoms – TIA or stroke (80.3%, 77.0%, respectively), compared to intermediate # 1 (68.3%) and less severe #0 and #4 (55.8% and 61.1% respectively) (p=0.001) (Table 1).

Next, we compared the baseline clinical characteristics in different clusters. We observed a non-random distribution of age (mean 67.2y SD:8.88, 69.1y SD:8.80, 70.4y SD:9.23, 67.5y SD: 8.80 and 68.9y SD: 8.30 in clusters #0, #1, #2, #3 and #4 respectively, p=0.019) and total cholesterol levels (mean 4,59 mmol/L SD:1.27, 4.48 mmol/L SD:1.24, 4.11 mmol/L SD:1.08, 4.58 mmol/L SD: 1.32 and 4.44 mmol/L SD: 1.11 in clusters #0, #1, #2, #3 and #4 respectively, p=0.050) (Table 1). The sex of patients, smoking status, diabetes, hypertension, body mass index, triglycerides, LDL, HDL, and CRP were not significantly different between the transcriptome-driven plaque types - suggesting that the molecular-based clusters cannot be directly approximated from basic clinical demographics.

### Evidence that genetic differences contribute to transcriptome-defined molecular plaque types

Atherosclerosis is a complex disease with a significant genetic component. Up now, GWASs identified over 163 independent genetic loci associated with atherosclerotic disease^14^. We hypothesized that the transcriptome-defined clustering reflects the genetic component of atherosclerotic disease and compared polygenic risk scores (PRS) for coronary artery disease (CAD)^15^. We observed a non-random distribution (p = 0.003) with the highest PRS in plaque type #3 (Fig. 3a). Suggesting that this plaque type is at least partially driven by genetic factors involved in CAD. The expression of genes genetically associated with CAD using gene-based testing (MAGMA^16^) showed a similar trend, with the highest expression of CAD-associated genes in plaque type #3 (Fig. 3b).

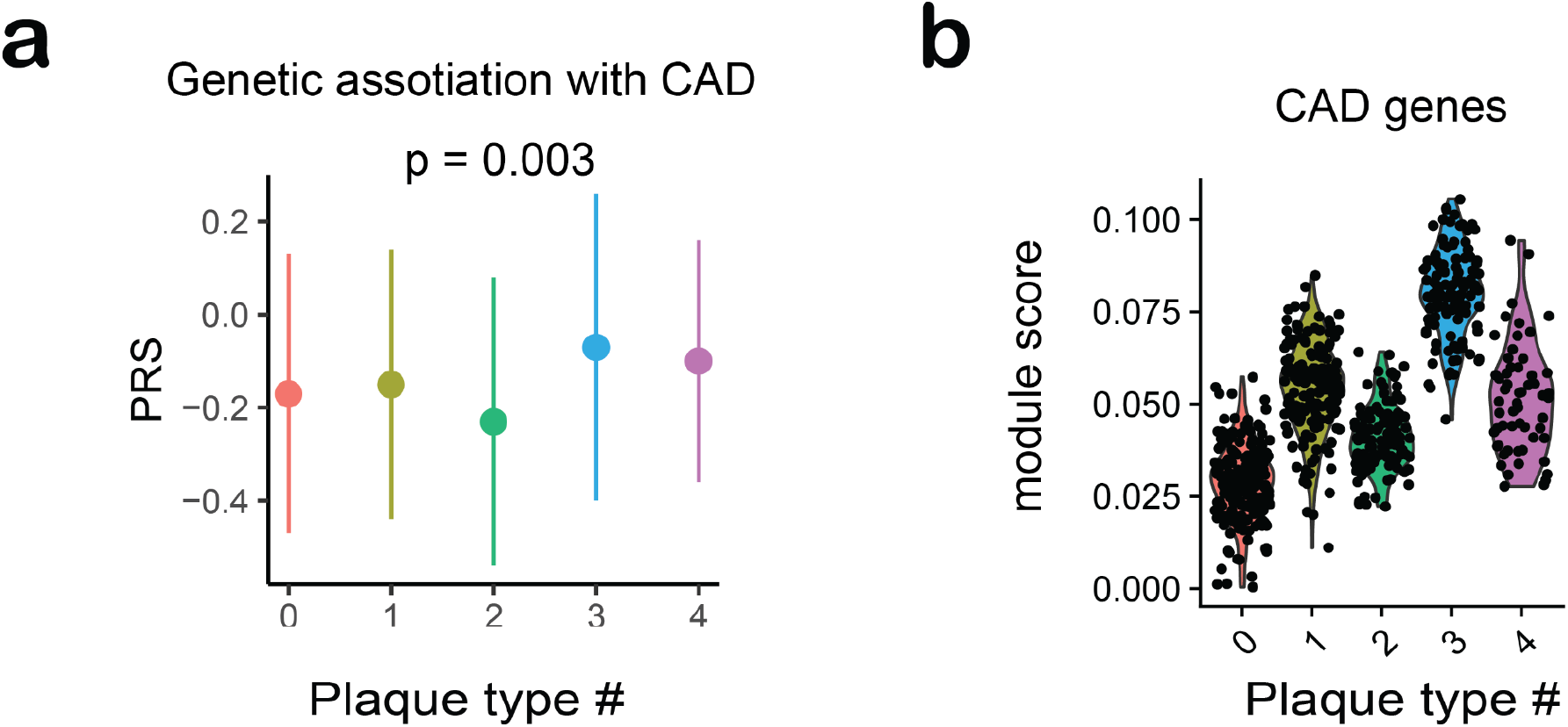

### Coronary artery lesions show similar transcriptomic clustering and associate with clinical presentation

To investigate whether plaques from other anatomical locations - the coronary arteries, present similar transcriptional profiles, we have repeated the clustering analysis on transcriptomics datasets derived from coronary arteries isolated from heart transplant recipient hearts^17^. We applied the same algorithm as for plaques from carotid arteries. In coronary artery segments, we were able to identify four distinct clusters (Fig. 4ab). Similar to carotids, the individual plaque types in coronary plaques show specific gene expression profiles, molecular pathways, and processes (Fig. 4e). Finally, the four clusters identified in coronary arteries differed in percentages of samples with underlying ischemia (18.2%, 54.9%, 6.5 % and 0.0%, respectively) (p < 0.001) (Fig. 4f). The cluster with the largest proportion of ischemic samples also showed increased expression of genes involved in ECM organization, neutrophil degranulation, TGF-beta signaling, and glycolysis. Notably, these are the same pathways associated with clinical symptoms in bulk transcriptomic analyses of carotid plaques in plaque type #3 (Fig. 1d).

**Figure 4.**
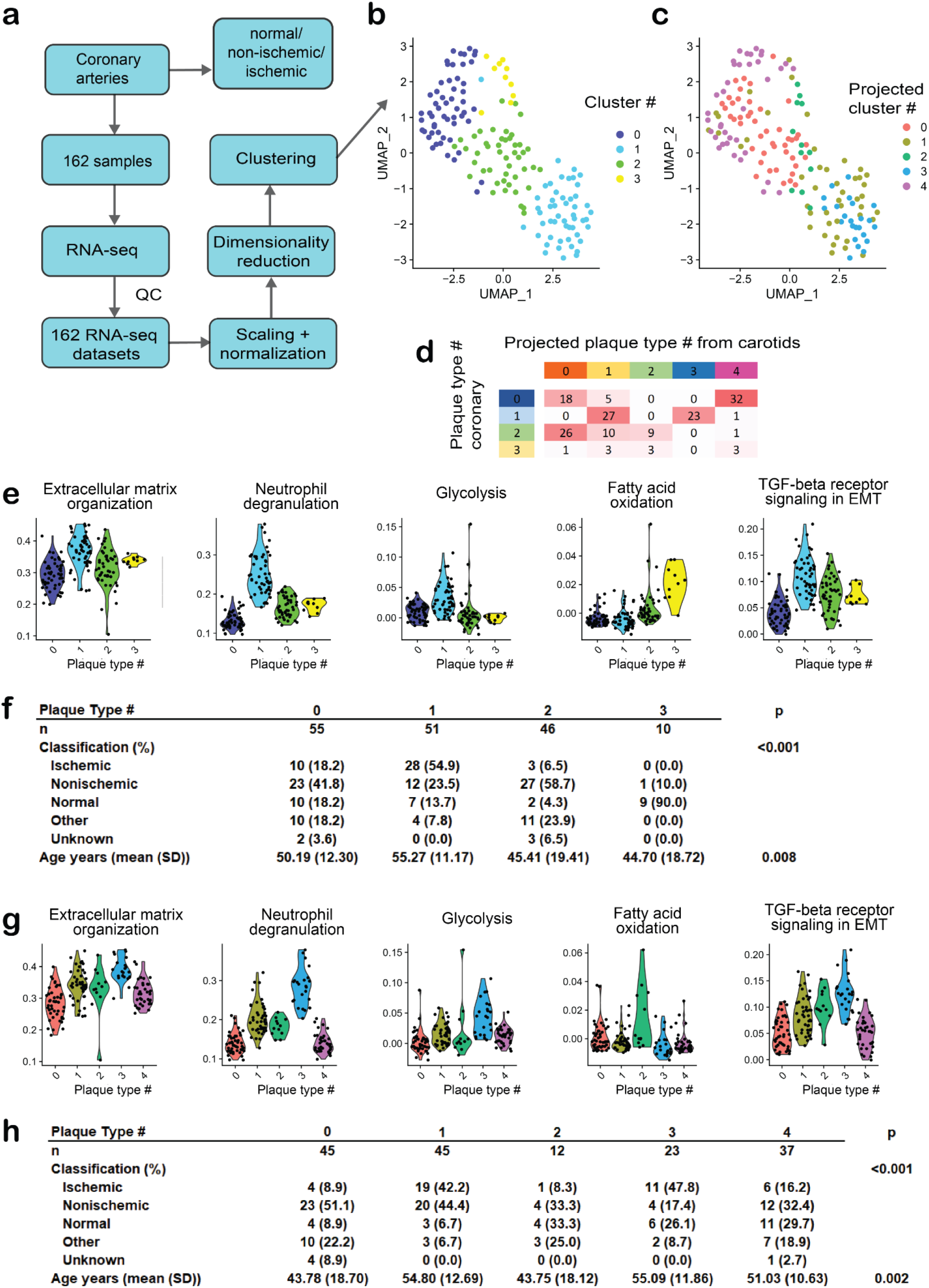
Unsupervised clustering of coronary artery samples based on transcriptomics data. a) A schematic workflow of the clustering analysis. b) UMAP projection of the 162 coronary samples based on RNA-seq dataset. The color indicates the cluster corresponding to the plaque type from the SNN modularity optimization-based clustering algorithm. c) UMAP projection of the 162 coronary samples based on RNA-seq dataset. The color indicates the projected plaque type identity from the carotid dataset. d) Correspondence of clusters derived from coronary data and projected clusters from carotids. Numbers in the table indicate sample counts in the intersection of the corresponding clusters. e) Module scores of genes annotated to selected molecular pathways in individual plaque clusters derived from coronary arteries. f) Distribution of clinical status and age of sample donors in coronary clusters. g) Module scores of genes annotated to selected molecular pathways in individual plaque clusters projected from carotid arteries. h) Distribution of clinical status and age of coronary artery sample donors in projected carotid clusters.

Since the coronary samples comprise the entire artery, including adventitial tissue, the four clusters identified in coronaries cannot be directly compared to the clusters from carotid arteries. Therefore, we aimed to identify the most similar carotid sample for each sample from coronary arteries. We have first employed the anchor-based data integration algorithm, which allows merging datasets with different confounders^18^. Next, we identified the best matching carotid sample to each coronary using the Pearson correlation (Fig. 4cd) and assigned the corresponding clusters.

The samples assigned to the same clusters again show the specific molecular pathway activity (like neutrophil degranulation or glycolysis) (Fig. 4g) and correlated with clinical manifestation (Fig. 4h). The best matching coronary samples for the patients detected in the symptomatic carotid plaque type #3 were observed in coronary cluster#1, which is the plaque type with the highest prevalence of ischemic cardiac disease. This suggests that the plaque types identified in this study and their connection to the plaque’s clinical manifestation translate to other cohorts and anatomical localizations.

### Circulating biomarkers correlate with transcriptomic clusters

A novel description and categorization of ‘vulnerable’ plaque types that goes beyond the scope of histopathological phenotyping is of great value for the scientific community in the field of vascular biology. However, for this novel type of patient and plaque stratification should be translated into tangible results – like circulating biomarkers applicable in a clinical setting. For clinical translation, we analyzed cardiovascular OLINK biomarker panels in available blood samples of 208 patients, searching for circulating biomarkers that associate the different transcriptomic clusters.

Out of 264 blood-derived biomarkers, we found that 10 were nominally associated with patho-histological plaque type (atheromatous, fibroatheromatous, or atheromatous) and 10 with the presence of calcifications (Fig. 5a). However, 28 circulating biomarkers showed significantly different levels among transcriptomic plaque clusters (Fig. 5a). Notably, the *PDGF subunit B* (p=0.005) showed the highest plasma levels in patients from plaque type #0. This growth factor is a potent activator of proliferation in cells of mesenchymal origin, and its receptor was recently shown to induce SMC-to- myofibroblast transitions in an aerobic glycolysis-dependent manner^19^ (Fig 5b). Another marker *SLAMF7* showed the highest plasma levels in patients with plaque type #2. Previously it was shown that the depletion of SLAMF7 in plaque-derived macrophages induced a suppressed secretion of proinflammatory cytokines, and inhibited proliferation of vascular smooth muscle cells^20^. This altogether suggests that the presence of distinct transcriptome-defined plaque types can be proxied by relevant circulating biomarkers.

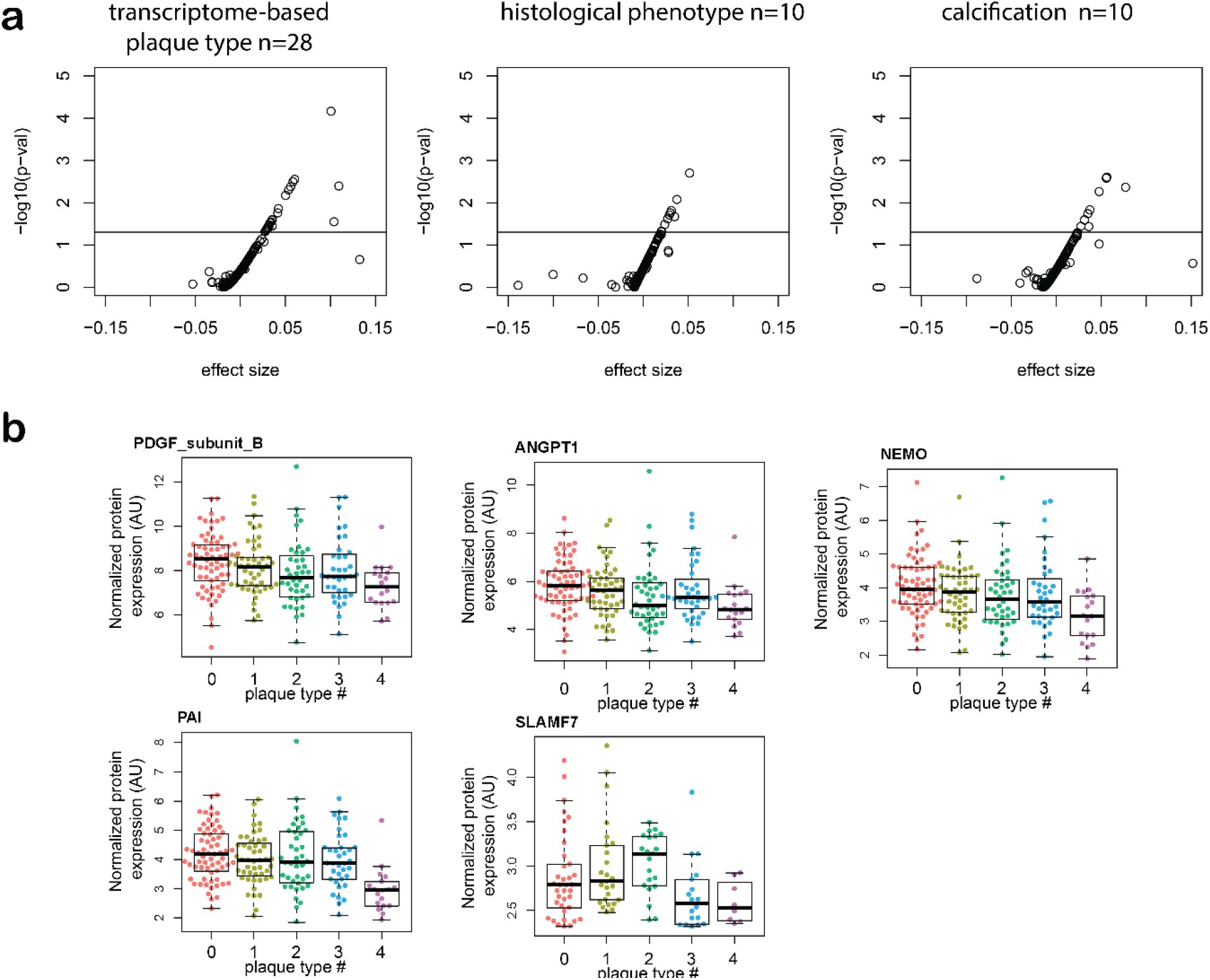

## Discussion

In the present study, using the unsupervised clustering, we describe five transcriptomic clusters in carotid plaques that relate to biological pathways and clinical presentation. Pathways that appeared overrepresented in a plaque type with more vulnerable patients pointed towards, among others, neutrophil degranulation, matrix organization, and glycolysis.

Atherosclerosis is a chronic lipid and inflammation-driven disease that develops and progresses over decades^21, 22^. The natural history of atherosclerotic plaque progression and stabilization is mainly unknown, and strikingly, the currently applied plaque description of the ‘vulnerable plaque’ has been initially based on pathology studies that reflect the worst cases, those who died^1, 2, 23^. The pathology-based concepts of the ‘vulnerable plaque’ have provided invaluable insights that have dominated vascular biology research for decades. Pathological observations have also shown the heterogeneity of vascular lesions that can lead to an adverse event. Plaque rupture has long been considered the pathological substrate leading to an acute thrombotic occlusion of the artery and described the lipid-rich, atheromatous, plaque with a thin fibrous cap with local infiltration of inflammatory cells that cause proteolytic activity and degradation of the stabilizing stimulate extracellular matrix. The mechanisms that underlie superficial erosion, a cause of coronary thrombosis distinct from plaque rupture, have garnered increasing interest^23–25^. In an era of improved control of traditional risk factors, plaque erosion assumes greater clinical importance^26^. Besides, the widely available analyses of genomic variants revealed novel and unexplored causal genes associated with the occurrence of acute atherosclerotic cardiovascular events^27^. The observation of thrombotic occlusion on an unruptured plaque^24^ and the role of microcalcifications^28^ destabilizing the cap made it clear that there is not one unambiguous description characterizing all unstable atherosclerotic lesions. The limitations of the current conceptual description of the ‘vulnerable plaque’ are also shown by the hallmark PROSPECT study that examined the natural history of advanced atherosclerotic lesions by serial intravascular imaging. There it was demonstrated that the predictive value of a thin capped fibro-atheromatous plaque is only 5% for the development of an adverse myocardial event. This increases to 20% if the presence of expansive remodeling and plaque cross-sectional area is taken into account^29, 30^.

The acceleration of RNA sequencing efforts of human atherosclerotic lesions provides an opportunity to fine-tune the description of the plaque at risk for a thrombotic event^12, 31, 32^.

### Pathology and transcriptome-based plaque characterization

Our transcriptomic data indicate the existence of plaque types that partly overlap with the pathological characterization and are associated with clinical symptoms of ischemic stroke and myocardial ischemia. Our analyzes also provide insight into cell-specific processes that can play a role in the destabilization of the plaque. Surprisingly, carotid plaques in plaque type #3, with the most severe symptoms and with the best match with symptomatic coronary plaques, showed enrichment of pathways and RNA-based cell types that are traditionally categorized in two distinct plaque categories. This plaque type revealed an abundance of the smooth muscle cell and macrophage-specific genes, cell types that in pathology and animal experimental categorizations are often considered as “stabilizing” and “destabilizing”, respectively. Our transcriptomic analyses confirm the existence of a complex landscape of atherosclerotic lesion phenotypes that associate with clinical symptoms. The inferences regarding the risk of clinical thrombotic events based on pathological therefore merit careful (re)consideration. Projection of the coronary transcriptome clusters onto the carotid cluster analysis showed substantial overlap.

### Transcriptomic clustering points to plaque molecular phenotypes that associate with increased risk for ischemic events

In both carotid and coronary plaques, there was a strong distinction between clusters in the expression of genes annotated to different molecular pathways.

Pathways indicating neutrophil activation, glycolysis, extracellular matrix organization, and iron uptake and transport were strongly over-represented in both arteries in the same cluster that was also associated with clinical events. This plaque type showed an overrepresentation of *CXCL12*, *CD14*, *C1QA*, *CD63*, *CD74*, *APOE*; genes that are often observed in plaque-derived macrophages. Indeed, deconvolution of the bulk sequencing data, using the cell-based transcripts from the single-cell sequencing, demonstrated that a significant proportion of the transcripts in this plaque type pointed to monocyte/macrophage lineages (Fig. 2cd).

The role of neutrophil activation in plaque stabilization has long been underestimated, but the discovery that neutrophil extracellular traps (NETs) may have a causal role in plaque destabilization confirms a potential role of genes indicating neutrophil activation^33–35^. Using in vivo and in vitro assays, Soehnlein and colleagues showed that activated SMCs in atherosclerotic plaques release chemotactic factors that attract neutrophils and trigger the release of NETs containing histone H4, which has cytotoxic effects on SMCs^36^. In addition, it has been shown that neutrophil microvesicles accumulate at disease-prone regions of arteries exposed to disturbed flow patterns and promote vascular inflammation and atherosclerosis in a murine model^37^.

The role of metabolic pathways such as glycolysis^38^, fatty acid oxidation, and tryptophan catabolism may point to phenotype switching of vascular cells^19^. The differentiation of a contractile to a synthetic myofibriblast-like phenotype in smooth muscle cells is driven by switching from oxidative to glycolytic metabolism^39^. In endothelial cells, glycolysis is essential for ATP production and sprouting of vessels since the loss of the glycolytic activator *PFKFB3* in ECs impairs vessel formation^40^. In addition, enhanced glycolysis has been recognized as a critical role in initiating endothelial or epithelial to mesenchymal transition (EMT) progression^41^. Smooth muscle cell and endothelial cell differentiation are mediated by local inflammation. The increased activity of the glycolysis pathway may thus relate to an active state of cell types that differentiate and accommodate to an unstable (inflammatory) environment.

The role of iron uptake in atherosclerosis has been debated and is considered multifaceted. Iron can cause oxidative damage by lipid peroxidation, and oxidized lipoprotein can then be taken up by the LDL receptor on macrophages leading to their development into foam cells (reviewed in ^42^). On the other hand, *CD163*+ alternative macrophages engulfing the hemoglobin-haptoglobin complexes (HH) were shown to augments hyaluronan synthesis in vascular SMCs and preventing vascular calcification^43^.

Genes overexpressed in the plaque transcriptomic type that predicted stroke and cardiac ischemia (like *CD14*) support the involvement of inflammation in this high-risk plaque. *CXCL12* is one of the genes that is also recognized in a GWAS for coronary artery disease^44^. *CD63* is a protein that is mainly associated with the membrane of intracellular vesicles and is also used as a marker for blood platelet activation^45^ as well as circulating vesicles^46^ and functionally plays a role in signal transduction. *C1QA* is considered to play a role in removing apoptotic cells via efferocytosis, and its presence reduces the atherosclerotic plaque size in animal models^47^.

### Clinical value of transcriptomic-based plaque characterization

The pathology-based description of the ‘vulnerable plaque’ has proven crucial value for understanding the progression and complications of atherosclerotic disease in experimental research^48^. In addition, the clinical utility of numerous vascular imaging modalities is based on the well-known definition of thin cap fibroatheroma. The discovery and evaluation of pharmaceutical treatments rely on animal models that apply the pathology-based surrogate measures of destabilizing atherosclerotic disease. Our observations provide evidence that transcriptomic profiling and subsequent clustering of human plaques may have substantial added value in searching for suitable drug targets. These observations that significantly fine-tune the phenotype of the destabilizing plaque can be translated to animal models and cell culture systems used in drug-related research.

There is an unmet need for biomarkers associated with the presence of plaques at risk for a thrombotic ischemic event. Proteins can today be measured with high accuracy using multiplex methods^49^. We feel that the observed enrichment of biomarkers associated with different transcriptomic plaque clusters is promising. Our sample size with combined plaque transcriptomic and clinical data does not allow strong inferences and surely requires verification. Our data do show that non-stratified biomarker analyses in large pooled patient samples may mask subgroups in whom biomarker profiles could be predictive. For example, clusters #0 and #4 both encompassed patients with mild symptoms but had different associations with biomarker profiles.

In summary, our study shows that deciphering the transcriptomic profile of atherosclerotic lesions results in an updated description of the ‘vulnerable plaque’ with clinical relevance. Our results also demonstrate that transcriptomic analyses can contribute to assessing lesion phenotypes that are predisposed to a thrombotic event and potentially reveal underlying pathogenetic mechanisms.

## Methods

### Carotid plaque samples

The Athero-Express Biobank (AE) includes patients undergoing carotid endarterectomy, of which the study design has been published before^50, 51^. The AE study is an ongoing biobank, and extensive baseline characteristics, blood samples, and atherosclerotic plaque specimens are collected. Clinical data were obtained from patient files and through standardized questionnaires. The indication for CEA was based on the recommendations from the Asymptomatic Carotid Surgery Trial (ACST) for asymptomatic patients and the European Carotid Surgery Trial (ECST) and North American Symptomatic Carotid Endarterectomy Trial for symptomatic patients (NASCET). Indications for CEA were evaluated by a multidisciplinary vascular team. The removal of atherosclerotic plaques was performed by a team of experienced surgeons, and standardized treatment protocols were applied. All patients were examined by a neurologist for assessment of their preoperative neurologic status. For this study, subsequent patients were included who underwent carotid endarterectomy between 2002 and 2016 and of whom genotyping data were available. The performed study is in line with the Declaration of Helsinki and informed consent was provided by all study participants after the approval for this study by medical ethical committees of the different hospitals was obtained.

### Baseline characteristics

Baseline data were obtained by chart review and from extensive questionnaires completed by the participating patients that included questions on the history of cardiovascular disease, cardiovascular risk factors (smoking, hypertension, diabetes), and use of medication. Presenting symptoms and duplex stenosis were retrieved from patient charts. Symptom categories were “asymptomatic”, defined as not having ipsilateral cerebrovascular symptoms in the previous 6 months; “ocular” - amaurosis fugax, defined as ipsilateral mono-ocular blindness of acute onset lasting <24 hours; cerebral “transient ischemic attack” (TIA), defined as the ipsilateral focal neurologic deficit of acute onset lasting <24 hours; and ipsilateral “stroke”. Lipid spectra were determined in blood specimens drawn at baseline.

### Follow up

All patients answered a questionnaire 1, 2, and 3 years after the carotid endarterectomy. In case an adverse event was reported or suspected, the referring hospital or general practitioner was approached for additional medical information. The primary outcome was defined as a composite of endpoints including, any death of vascular origin (fatal stroke, fatal myocardial infarction, sudden death, and other vascular death), non-fatal stroke (either ischemic or hemorrhagic) or non-fatal myocardial infarction, and any arterial vascular intervention that had not already been planned at the time of inclusion (e.g. carotid surgery or angioplasty, coronary artery bypass, percutaneous coronary artery intervention, peripheral vascular surgery, or angioplasty).

### Sample handling

The atherosclerotic plaques were transported to the laboratory and processed immediately after the surgical removal. An experienced technician identified the culprit lesion, which is defined as the segment with the smallest lumen. In case of doubt, the segment with the largest plaque diameter was selected. The segment with the culprit lesion was then prepared and stored in 4% formaldehyde, decalcified, and embedded in paraffin for histological analysis. The rest of the plaque was snap-frozen using liquid nitrogen and stored at -80 degrees Celsius.

### Histological assessment

The assessment was performed according to a previously validated protocol, which was described in detail before^52, 53^. In brief, cross-sections of the culprit lesion are stained and quantified for each patient. A hematoxylin-eosin (HE) staining was performed for the assessment of calcification, picrosirius red for collagen and alpha-smooth muscle actin for smooth muscle cells. *CD68* was stained to identify macrophages. The location of SMCs was evaluated as “mainly basal”, “homogeneous,” and” mainly luminal”. The location of macrophages was evaluated as “basal”, “homogeneous”, ”luminal”, or “no macrophages”. Plaque characteristics were scored semi-quantitatively at 40x magnification and grouped into no, minor, moderate and heavy staining. In the present study, these categories were binned into no/minor staining and moderate/heavy staining. Immunohistochemical staining for *CD34* was performed to assess vessel density. Plaque microvessels were quantified in three hotspots and subsequently, the average number of vessels per hotspot was calculated. Picrosirius red in combination with elastic-Van Gieson, HE and polarized light was used to visualize the lipid core. The lipid content of the plaque was estimated as a percentage of the total plaque area, with a cutoff at 10% and 40% for carotid plaques. Plaques with<10%, 10-40% and >40% fat were categorized as fibrous, fibro-atheromatous and atheromatous, respectively. The presence of plaque hemorrhage was determined using HE staining, fibrin staining and glycophorin staining. All plaque characteristics were scored and quantified with good intra- and interobserver reproducibility by two independent observers^52^.

### RNA isolation and library preparation

A total of 700 segments were selected from patients who were included in the study between 2002 and 2016. The RNA isolated from the archived advanced atherosclerotic lesion is fragmented (Extended Data figure 2a). We have, therefore, tested four different library preparation strategies (Extended Data figure 2bc): CEL-seq2^54^, QIAseq (QIAseq Stranded Total RNA Lib Kit, Qiagen), NEXTflex (NEXTflex Rapid Directional RNA-Seq Kit, Bioo Scientific) and SMARTer (SMARTer® Stranded RNA-Seq Kit, Takara) using the manufacturer’s or author’s recommendations. We have ultimately employed the CEL-seq2 method^54^. CEL- seq2 yielded the highest mappability reads to the annotated genes compared to other library preparation protocols (Extended Data figure 2bc). The methodology captures 3’-end of polyadenylated RNA species and includes unique molecular identifiers (UMIs), which allow direct counting of unique RNA molecules in each sample. 50ng of total RNA was precipitated using isopropanol and washed with 75% ethanol. After removing ethanol and air-drying the pellet, primer mix containing 5ng primer per reaction was added, initiating primer annealing at 65°C for 5min. Subsequent RT reaction was performed; first strand reaction for 1h at 42°C, heat-inactivated for 10m at 70°C, second strand reaction for 2h at 16°C, and then put on ice until proceeding to sample pooling. The primer used for this initial reverse-transcription (RT) reaction was designed as follows: an anchored polyT, a unique 6bp barcode, a unique molecular identifier (UMI) of 6bp, the 5’ Illumina adapter and a T7 promoter, as described. Each sample now contained its own unique barcode due to the primer used in the RNA amplification, making it possible to pool together cDNA samples at 7 samples per pool. Complementary DNA (cDNA) was cleaned using AMPure XP beads (Beckman Coulter), washed with 80% ethanol, and resuspended in water before proceeding to the *in vitro* transcription (IVT) reaction (AM1334; Thermo-Fisher) incubated at 37°C for 13 hours. Next, primers were removed by treating with Exo-SAP (Affymetrix, Thermo-Fisher) and amplified RNA (aRNA) was fragmented and then cleaned with RNAClean XP (Beckman-Coulter), washed with 70% ethanol, air-dried, and resuspended in water. After removing the beads using a magnetic stand, RNA yield and quality in the suspension were checked by Bioanalyzer (Agilent).

cDNA library construction was then initiated by performing an RT reaction using SuperScript II reverse transcriptase (Invitrogen/Thermo-Fisher) according to the manufacturer’s protocol, adding randomhexRT primer as a random primer. Next, PCR amplification was done with Phusion High-Fidelity PCR Master Mix with HF buffer (NEB, MA, USA) and a unique indexed RNA PCR primer (Illumina) per reaction, for a total of 11-15 cycles, depending on aRNA concentration, with 30 seconds elongation time. PCR products were cleaned twice with AMPure XP beads (Beckman Coulter). Library cDNA yield and quality were checked by Qubit fluorometric quantification (Thermo-Fisher) and Bioanalyzer (Agilent), respectively. Libraries were sequenced on the Illumina Nextseq500 platform; paired end, 2 x 75bp.

### Sequencing read mapping and quality filtering

Libraries were sequenced on the Illumina Nextseq500 platform; a high output paired-end run of 2 × 75 bp was performed (Utrecht Sequencing Facility). The reads were demultiplexed and aligned to human cDNA reference (Ensembl 84) using the BWA (0.7.13) by calling ‘bwa aln’ with settings -B 6 -q 0 -n 0.00 -k 2 -l 200 -t 6 for R1 and -B 0 -q 0 -n 0.04 -k 2 -l 200 -t 6 for R2, ‘bwa sampe’ with settings -n 100 -N 100. Multiple reads mapping to the same gene with the same unique molecular identifier (UMI, 6bp long) were counted as a single read. The raw read counts were corrected for UMI sampling (corrected_count=-4096*(ln(1- (raw_count/4096)))), normalized for sequencing depth and quantile normalized. We have detected a median of 19.501 (SD = 5.874) genes per sample with at least one unique read (Extended Data figure 2d) and discarded samples (n=46) with less than 9000 detected genes from further analysis (Extended Data figure 2e and Fig 1a). For all the subsequent analyses, we have excluded all the ribosomal genes and used only the protein-coding genes with annotated HGCN names.

### Clustering of transcriptomics datasets

Clustering of datasets from carotid arteries was based on the first 12 principal components (PCs) calculated using 5000 most variable genes from the normalized gene expression data. We used the shared nearest neighbor (SNN) modularity optimization-based clustering algorithm^55^ implemented in the Seurat package^18^ (core scripts can be found at https://github.com/CirculatoryHealth/PlaqueCluster). 162 transcriptomics data sets from coronary arteries were clustered separately in the same way.

### Pathway analysis

Pathway analysis was performed using the “ReactomePA” R/Bioconductor package^56^. Module score were calculated using the Seurat’s AddModuleScore() function.

### Baseline characteristics tables

Baseline characteristics tables were produced using R’s “tableone” package with default settings. Statistical significance of differences between groups was tested using the chi-square test for categorical variables and one-way analysis of variance (ANOVA) for continuous variables (with equal variance assumption, i.e., regular ANOVA).

### Cell Deconvolution Analysis

We deconvoluted the gene expression of 632 advanced carotid plaques into cell composition matrices using the MUSIC deconvolution method with default parameter^11^ based on a single cell RNAseq data from advanced carotidatherosclerotic plaques^12^. We merged all the sub-cell types into 7 cell types (‘CD14+CD68+ Macrophages’,’CD34+ Endothelial Cells’,’CD3+CD4+ T Cells’,’CD3+CD8+ T Cells’,’CD79A+ B-Cells’,’KIT+ Mast Cells’,’ACTA2+ Smooth Muscle Cells) and removed mixed cells.

### Human coronary artery tissue procurement

Ischemic human coronary artery tissue biospecimens were obtained at Stanford University from diseased heart transplant donors consenting for research studies. Hearts were arrested in cardioplegic solution and transported on ice prior to dissecting proximal coronary artery segments from main branches of left anterior descending, circumflex or right coronary arteries. Epicardial and perivascular adipose was trimmed on ice, rinsed in cold phosphate-buffered saline, and rapidly frozen in liquid nitrogen, and stored at -80C. Normal human coronary artery tissue biospecimens were also obtained at Stanford University from non-diseased donor hearts rejected for orthotopic heart transplantation processed following the same protocol as hearts for transplant. Tissues were de-identified and clinical and histopathology information was used to classify ischemic and non-ischemic arteries. All normal arteries originated from hearts with a left ventricular ejection fraction (LVEF) greater than 50%. Frozen tissues were transferred to the University of Virginia through a material transfer agreement and Institutional Review Board-approved protocols.

### RNA Extraction, QC, library construction and sequencing of coronary samples

Total RNA was extracted from frozen coronary artery segments using the Qiagen miRNeasy Mini RNA Extraction kit (catalog #217004). Approximately 50 mg of frozen tissue was pulverized using a mortar and pestle under liquid nitrogen. Tissue powder was then further homogenized in Qiazol lysis buffer using stainless steel beads in a Bullet Blender (Next Advance) homogenizer, followed by column-based purification. RNA concentration was determined using Qubit 3.0 and RNA quality was determined using Agilent 4200 TapeStation. Samples with RNA Integrity Number (RIN) greater than 5.5 and Illumina DV200 values greater than 75 were included for library construction. Total RNA libraries were constructed using the Illumina TruSeq Stranded Total RNA Gold kit (catalog #20020599) and barcoded using Illumina TruSeq RNA unique dual indexes (catalog # 20022371). After re-evaluating library-quality using TapeStation, individually barcoded libraries were sent to Novogene for next-generation sequencing. After passing additional QC, libraries were multiplexed and subjected to paired-end 150 bp read sequencing on an Illumina NovaSeq S4 Flowcell to a median depth of 100 million total reads (>30 G) per library.

### RNA-seq processing and analysis of coronary samples

The raw passed filter sequencing reads obtained from Novogene were demultiplexed using the bcl2fastq script. The quality of the reads was assessed using FASTQC and the adapter sequences were trimmed using trimgalore. Trimmed reads were aligned to the hg38 human reference genome using STAR v2.7.3a according to the GATK Best Practices for RNA-seq. To increase mapping efficiency and sensitivity, novel splice junctions discovered in a first alignment pass with high stringency were used as annotation in a second pass to permit lower stringency alignment and therefore increase sensitivity. PCR duplicates were marked using Picard and WASP was used to filter reads prone to mapping bias. Total read counts and RPKM were calculated with RNA-SeQC v1.1.8 using default parameters and additional flags “-n 1000 - noDoC -strictMode” and GENCODE v30 reference annotation.

### Projection of coronary artery datasets with carotid based clusters

To integrate two heterogeneous datasets, we have first used the anchor-based data integration algorithm^18^. Next, we have created a pairwise correlation matrix between individual samples using the Pearson correlation. Then for each coronary sample, we have assigned the cluster identity of the closest (best positively correlated) carotid sample.

### Measurement of circulating biomarkers

In 208 selected patients from AtheroExpress cohort, we used a commercially available multiplex proximity extension assay^57^ from Olink proteomics AB platform (Uppsala, Sweden) to measure 264 proteins using the Olink® Cardiovascular II, Olink® Cardiovascular III, and Olink® Cardiometabolic panels. These panels were selected for their known associations with CV disease. Proteins are expressed on a log2- scale as normalized protein expression (NPX) values. Patients were randomly distributed across plates.

### Genotyping, imputation and weighted polygenic scores calculation

#### DNA isolation and genotyping

We genotyped the AE in three separate but consecutive experiments^58^. The DNA was extracted from EDTA blood or (when no blood was available) plaque samples using in-house validated protocols and genotyped in 3 batches (Athero-Express Genomics Studies). For the Athero-Express Genomics Study 1 (AEGS1), included between 2002 and 2007, were genotyped (440,763 markers) using the Affymetrix Genome-Wide Human SNP Array 5.0 (SNP5) chip (Affymetrix Inc., Santa Clara, CA, USA) at Eurofins Genomics, https://www.eurofinsgenomics.eu/). For the Athero-Express Genomics Study 2 (AEGS2), included between 2002 and 2013, were genotyped (587,351 markers) using the Affymetrix AxiomⓇ GW CEU 1 Array (AxM) at the Helmholtz Genome Analysis Center (https://www.helmholtz-muenchen.de/no_cache/gac/index.html). For the Athero-Express Genomics Study 3 (AEGS3), included between 2002 and 2016, were genotyped (693,931 markers) using the Illumina GSA MD v1 BeadArray (GSA) at Human Genomics Facility, HUGE-F (http://glimdna.org/index.html). All experiments were carried out according to OECD standards and as advised by the respective manufacturer. We used the genotyping calling algorithms as advised by Affymetrix (BRLMM-P for AEGS1 and AxiomGT1 for AEGS2) and Illumina (Illumina GenomeStudio For AEGS3).

#### Quality control after genotyping

After genotype calling, we adhered to community standard quality control and assurance (QCA) procedures of the genotype data from AEGS1, AEGS2, and AEGS3^59^. Samples with low average genotype calling and sex discrepancies (compared to the clinical data available) were excluded. The data was further filtered on: 1) individual (sample) call rate > 97%, 2) SNP call rate > 97%, 3) minor allele frequencies (MAF) > 3%, 4) average heterozygosity rate ± 3.0 cs.d., 5) relatedness (pi-hat > 0.20), 6) Hardy– Weinberg Equilibrium (HWE p < 1.0×10^−3^, 7) Monomorphic SNPs (MAF< 1.0×10^−6^), and 8) deviation in the principal component analysis plot using 1000G phase 3 as reference (6 iterations ± 3s.d.).

#### Imputation

Before phasing using SHAPEIT2^60^, data was lifted to genome build b37 using the liftOver tool from UCSC (https://genome.ucsc.edu/cgi-bin/hgLiftOver). Finally, data were imputed with 1000G phase 3, version 5 and HRC release 1.1 as a reference using the Michigan Imputation Server (https://imputationserver.sph.umich.edu/). These results were further integrated using QCTOOL v2 (https://www.well.ox.ac.uk/~gav/qctool_v2/), where HRC imputed variants are given precedence over 1000G phase 3 imputed variants. After imputation, we compared the quality of the three AEGS datasets based on sample type (EDTA blood or plaque) and genotyping chip used. We checked identity-by-descent (IBD) within and between datasets to aid in sample mixups, duplicate sample use, and relatedness.

#### Weighted polygenic score calculation

We estimated the weighted polygenic cardiovascular disease susceptibility using the previously published polygenic risk score for coronary artery disease (MetaGRS) described before^61^. Briefly, the MetaGRS comprises 1,745,179 genetic variants with a minor allele frequency (MAF) > 0.1% associated with CAD and was constructed through meta-analysis of three genomic risk scores: GRS46K (comprising 46,000 cardiometabolic genetic variants), FDR202 (including 202 genetic variants associated with CAD at false discovery rate *p* < 0.05 in the recent GWAS CARDIoGRAMplusC4D), and the 1000Genomes genetic score also created with CARDIoGRAMplusC4D. The MetaGRS was internally and externally validated for the primary risk of prevalent and incident CAD in the UK Biobank^61^. We matched the 1.7 million variants from the MetaGRS to 1,742,593 variants in our data (2586 variants were not present in our data). Given that the median imputation quality was high (INFO = 0.978 [IQR 0.945–0.991]), and the variants included in the MetaGRS have MAF >0.1%, we did not further filter on imputation quality. Moreover, since we used the imputed genotype probabilities to calculate the MetaGRS, rather than the hard-coded genotypes, bias arising from imputation error, i.e., low imputation quality, will only reduce predictive accuracy. Thus, we calculated the weighted polygenic score (PGS) for each included patient in this study using PRSice-2^62^ as follows. To account for the imputation quality, we used the allelic dosages (*D*) estimated by IMPUTE2 based on the posterior genotype probabilities (*Pg*) for the B-allele (B) for the *i*^th^ variant. Thus:

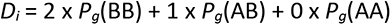

For each individual, we calculated the aggregate polygenic scores using the effect *D* of all modeled variants weighted by the effect size (β) of the *i*^th^ variant as given in the MetaGRS. Thus, for each individual *n*, the weighted PGS is the sum of the β of the *i*^th^ variant multiplied by the dosage *D* of that respective variant:

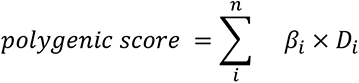

We standardized the PGS to mean-zero and unit-variance for each genotyping batch separately.

## Tables

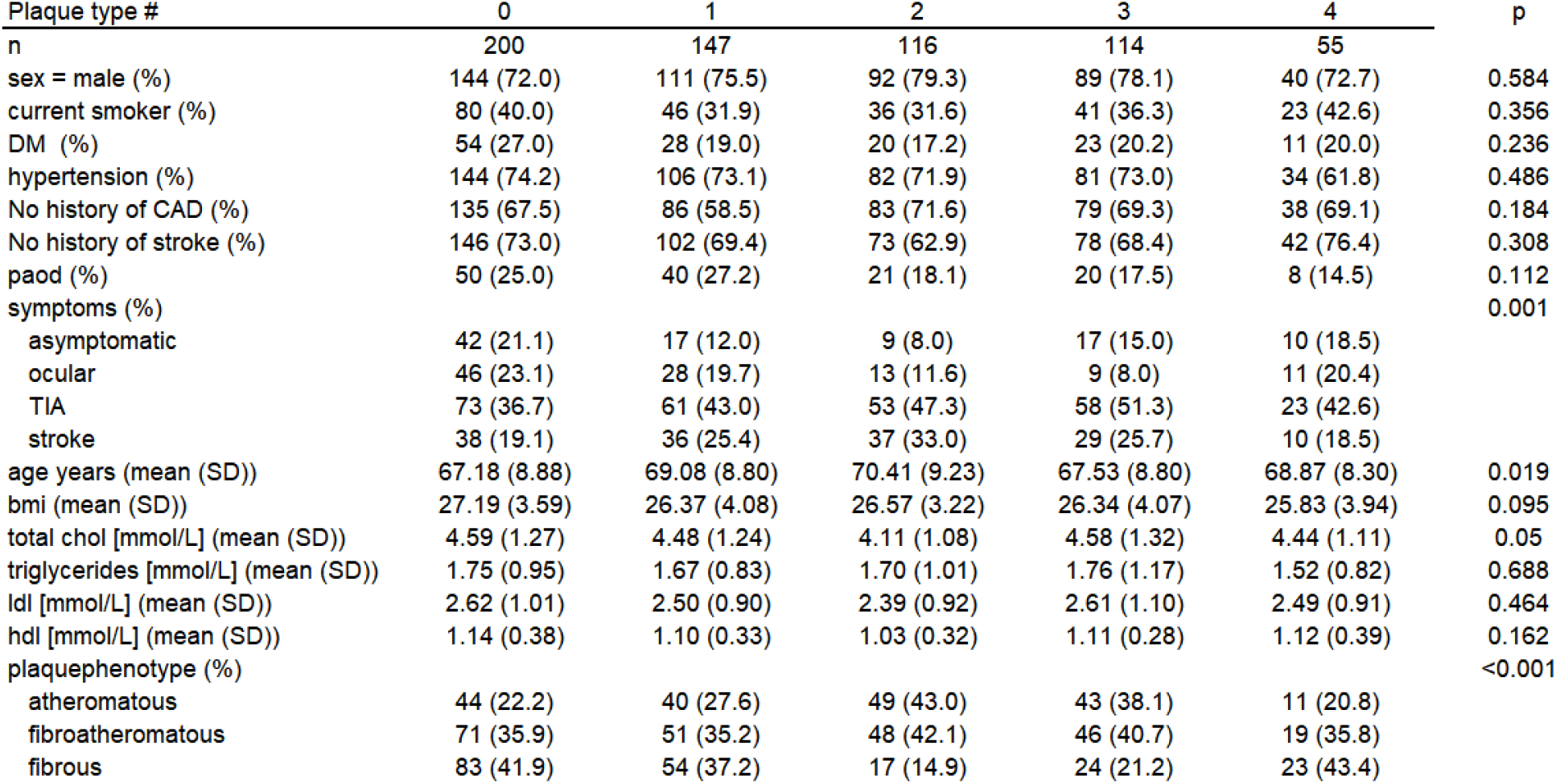

| Plaque type # | 0 | 1 | 2 | 3 | 4 | p |
| --- | --- | --- | --- | --- | --- | --- |
| n | 200 | 147 | 116 | 114 | 55 |  |
| sex = male (%) | 144 (72.0) | 111 (75.5) | 92 (79.3) | 89 (78.1) | 40 (72.7) | 0.584 |
| current smoker (%) | 80 (40.0) | 46 (31.9) | 36 (31.6) | 41 (36.3) | 23 (42.6) | 0.356 |
| DM (%) | 54 (27.0) | 28 (19.0) | 20 (17.2) | 23 (20.2) | 11 (20.0) | 0.236 |
| hypertension (%) | 144 (74.2) | 106 (73.1) | 82 (71.9) | 81 (73.0) | 34 (61.8) | 0.486 |
| No history of CAD (%) | 135 (67.5) | 86 (58.5) | 83 (71.6) | 79 (69.3) | 38 (69.1) | 0.184 |
| No history of stroke (%) | 146 (73.0) | 102 (69.4) | 73 (62.9) | 78 (68.4) | 42 (76.4) | 0.308 |
| paod (%) | 50 (25.0) | 40 (27.2) | 21 (18.1) | 20 (17.5) | 8 (14.5) | 0.112 |
| symptoms (%) |  |  |  |  |  | 0.001 |
| asymptomatic | 42 (21.1) | 17 (12.0) | 9 (8.0) | 17 (15.0) | 10 (18.5) |  |
| ocular | 46 (23.1) | 28 (19.7) | 13 (11.6) | 9 (8.0) | 11 (20.4) |  |
| TIA | 73 (36.7) | 61 (43.0) | 53 (47.3) | 58 (51.3) | 23 (42.6) |  |
| stroke | 38 (19.1) | 36 (25.4) | 37 (33.0) | 29 (25.7) | 10 (18.5) |  |
| age years (mean (SD)) | 67.18 (8.88) | 69.08 (8.80) | 70.41 (9.23) | 67.53 (8.80) | 68.87 (8.30) | 0.019 |
| bmi (mean (SD)) | 27.19 (3.59) | 26.37 (4.08) | 26.57 (3.22) | 26.34 (4.07) | 25.83 (3.94) | 0.095 |
| total chol [mmol/L] (mean (SD)) | 4.59 (1.27) | 4.48 (1.24) | 4.11 (1.08) | 4.58 (1.32) | 4.44 (1.11) | 0.05 |
| triglycerides [mmol/L] (mean (SD)) | 1.75 (0.95) | 1.67 (0.83) | 1.70 (1.01) | 1.76 (1.17) | 1.52 (0.82) | 0.688 |
| ldl [mmol/L] (mean (SD)) | 2.62 (1.01) | 2.50 (0.90) | 2.39 (0.92) | 2.61 (1.10) | 2.49 (0.91) | 0.464 |
| hdl [mmol/L] (mean (SD)) | 1.14 (0.38) | 1.10 (0.33) | 1.03 (0.32) | 1.11 (0.28) | 1.12 (0.39) | 0.162 |
| plaquephenotype (%) |  |  |  |  |  | <0.001 |
| atheromatous | 44 (22.2) | 40 (27.6) | 49 (43.0) | 43 (38.1) | 11 (20.8) |  |
| fibroatheromatous | 71 (35.9) | 51 (35.2) | 48 (42.1) | 46 (40.7) | 19 (35.8) |  |
| fibrous | 83 (41.9) | 54 (37.2) | 17 (14.9) | 24 (21.2) | 23 (43.4) |  |

## Data Availability

The data are not publicly available due to research participant privacy/consent

## Acknowledgements

The authors would like to thank the Utrecht Sequencing Facility for continuous support and patience. This work was supported by The Dutch Heart Foundation (CVON2017-20: Generating the best evidence-based pharmaceutical targets and drugs for atherosclerosis [GENIUS II] to G. Pasterkamp, S.W. van der Laan); Fondation Leducq (Transatlantic Network Grant PlaqOmics to N. J. Leeper, M. Civelek. G. K. Owens, A. V. Finn, Clint. L. Miller and G. Pasterkamp); EU 755320 Taxinomisis grant (E. Pavlos, E. Andreakos, G.J. de Borst, A. Boltjes, G. Pasterkamp). We acknowledge the European Research Area Network on Cardiovascular Diseases (ERA-CVD, grant number 01KL1802 to F. W. Asselbrgs, S.W. van der Laan, G. Pasterkamp); the ERA-Endless consortium (Dutch Heart Foundation, grant number 2017/T099 to H.M. den Ruijter and G. Pasterkamp), European Research Council (ERC) consolidator grant (grant number 866478 UCARE to H.M. den Ruijter). The authors would like to thank the Utrecht Sequencing Facility for continuous support and patience.

## Author contributions

MM AB analyzed and integrated the data. SWv/dL and JM provided PRS calculations. AB, SWv/dL, performed patient selection, randomization and sample handling. GJdeB performed carotid endarterectomy procedures. NAMv/dD, NL and EM tested library preparation strategies and processed coronary samples for sequencing. MACD, KHMP, LS, MPJW, JP provided, analyzed and interpreted single cell sequencing data. KC performed deconvolution of bulk RNA-seq data. JM, NT, FW, MCV recruited the patients coordinated by DPVdeK. DPVdeK, ESGS provided and coordinated Olink measurements. LS and SWv/dL performed MAGMA analysis. AB, NT, FW, DPVdeK, HMdenR, FWA, SWv/d Laan participated in conceptualization, data interpretation, and provided critical feedback on the article. EDB, RJGH, EP, EA, HS, GKO, CM, AVF, RV, NJL and MC participated in data interpretation, and provided critical feedback on the article. AWT, MDK, CJH and CLM recruited, processed and analyzed coronary samples. EA, FWA, SWv/dL, CLM, MM, GP provided funding. MM, CLM and GP participated in the conceptualization and supervision of the project and finalization of the article. MM prepared the figures. MM and GP drafted the manuscript. All authors provided feedback on the research, analyses, and article.

## Data availability

The data are not publicly available due to research participant privacy/consent.

## Code availability

The core scripts used for the analysis can be found at https://github.com/CirculatoryHealth/PlaqueCluster

