## Supplementary figures and table for "Transcriptomic-based clustering of advanced atherosclerotic plaques identifies subgroups of plaques with differential underlying biology that associate with clinical presentation"

**Supplementary Table 1** Baseline characteristics

| | |
|---|---|
| n | 632 |
| sex = male (%) | 476 (75.3) |
| current smoker (%) | 226 (36.2) |
| DM (%) | 136 (21.5) |
| hypertension (%) | 447 (72.2) |
| symptoms (%) | |
| asymptomatic | 95 (15.3) |
| ocular | 107 (17.3) |
| TIA | 150 (24.2) |
| stroke | 268 (43.2) |
| age years (mean (SD)) | 68.43 (8.92) |
| bmi (mean (SD)) | 26.61 (3.78) |
| totalchol [mmol/L] (mean (SD)) | 4.47 (1.23) |
| triglyceriden [mmol/L] (mean (SD)) | 1.71 (0.96) |
| ldl [mmol/L] (mean (SD)) | 2.54 (0.98) |
| hdl [mmol/L] (mean (SD)) | 1.11 (0.35) |
| plaquephenotype (%) | |
| atheromatous | 187 (30.0) |
| fibroatheromatous | 235 (37.7) |
| fibrous | 201 (32.3) |

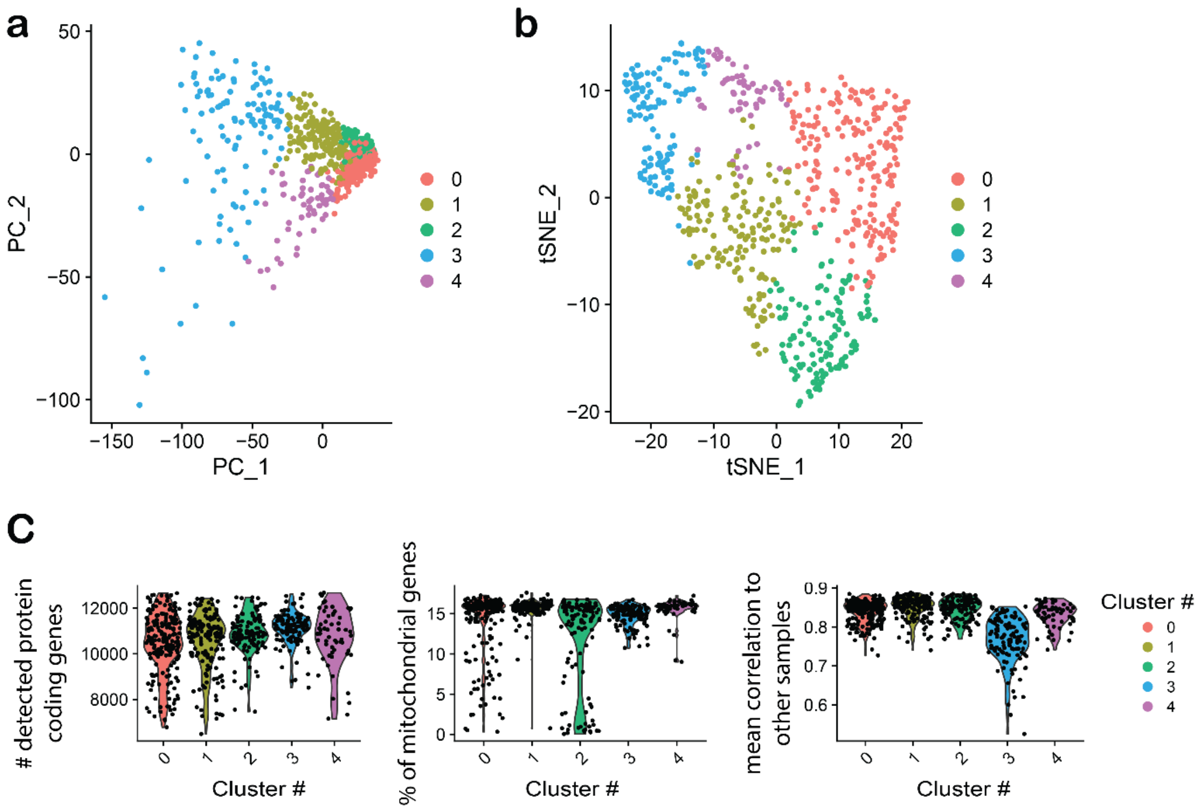


***Supplementary Figure 1 Unsupervised clustering of plaques based on transcriptomics data**. **a)** PCA plot and **b)** tSNE projection of the 654 plaque samples based on RNA-seq dataset. The color indicates the cluster corresponding to the plaque type cluster from the SNN modularity optimization based clustering algorithm. **c)** Distribution of non-ribosomal protein-coding genes with annotated HGNC name; reads mapping to mitochondrial genes and mean Pearson correlation of samples per cluster*

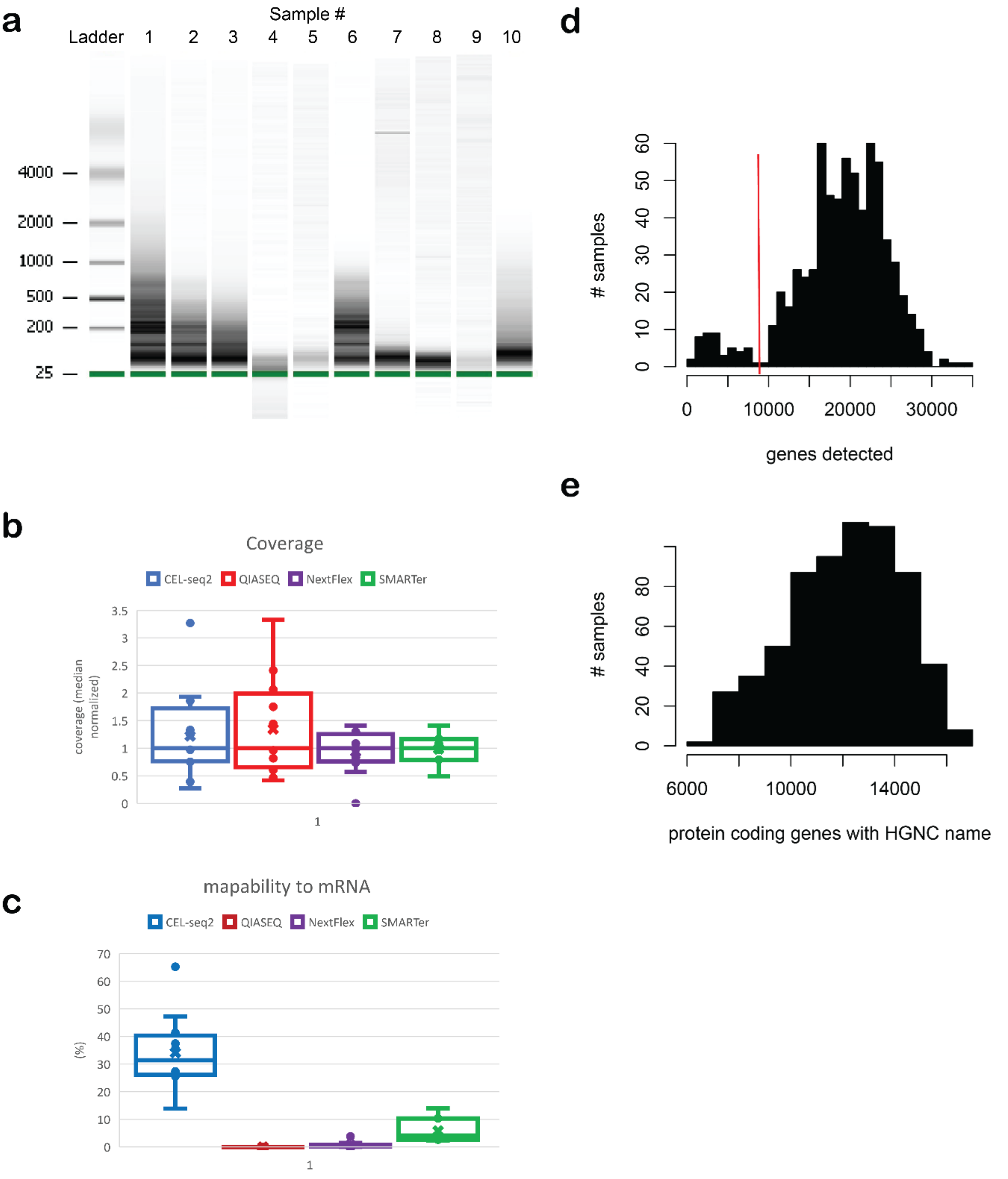


***Supplementary Figure 2. Plaque transcriptomics. a)*** *representative bioanalyzer profiles of total RNA isolated from 10 samples of advanced atherosclerotic lesions.* ***b)*** *Distribution of sequencing reads between samples using four different library preparation strategies.* ***c)*** *Percentage of sequenced reads mapped to annotated genes using four different library preparation strategies.* ***d****) Number of annotated genes identified per sample with at least one mapped read. Samples with less than 9000 genes were excluded from the analysis.* ***e)*** *Number of non-ribosomal protein-coding genes with annotated HGNC name per sample used in the analysis.*
